# Real-time fMRI neurofeedback training as a neurorehabilitation approach on depressive disorders: A systematic review of randomized control trials

**DOI:** 10.1101/2020.08.09.20159319

**Authors:** Pamela González Méndez, Ranganatha Sitaram, Jeffrey A. Stanley, Julio Rodiño Climent

## Abstract

**BACKGROUND:** Depressive disorders are a group of neuropsychiatric disorders that cause significant distress and impairment in social, occupational, and other important areas of functioning. In the last decade, Brain-Computer Interfaces based-neurofeedback training appears as an innovative therapy for this condition and other neuropsychiatric disorders, allowing to volitionally self-regulate brain activity and behavior. Up to date, non-invasive neurofeedback training have been built on different techniques, including EEG, NIRS and fMRI.

**OBJECTIVES:** This systematic review aims to evaluate the clinical application of fMRI neurofeedback training and its efficacy on treating depressive disorders. As a secondary objective, we intend to extract additional information on the neurofeedback training technique, in order to provide recommendations for future research.

**METHODS AND ANALYSIS:** The systematic review complies with the PRISMA guidelines and it was submitted to PROSPERO registration. We will only include randomized control trials assessing participants with a depressive disorder. The intervention of interest is real-time fMRI neurofeedback training, the comparison of interest will be placebo neurofeedback, another active non-neurofeedback control or no treatment. The primary outcome will be effects on behavior (symptomatology/disease severity reduction). The secondary outcomes will assess quality of life, acceptability and adverse effects. Finally, we will evaluate ‘other outcomes’ regarding brain MRI metrics (BOLD activation/connectivity), cognitive tasks performance, and physiology measures. At least two reviewers will independently select studies, extract data and assess the risk of bias. If methodologically possible, for primary and secondary outcomes, a meta-analysis will be performed and the data will be presented in summary tables of results using the GRADE approach.

**STRENGTH AND LIMITATIONS:** As the number of studies on neurofeedback is increasing every year, and better quality of evidence is available, this systematic review, will include only randomized control trials. To our knowledge, this is the first systematic review assessing randomized control trials on fMRI neurofeedback training as a neurorehabilitation approach on depressive disorders. The main limitation of this systematic review might arise from the low number of extant RCTs.

## INTRODUCTION

Depression is a medical condition different from usual mood fluctuations and emotional responses to challenges in everyday life, which is accompanied by somatic and cognitive changes that signifi-cantly affect the individual’s capacity to function. Its symptoms cause significant distress or impairment in social, occupational, or other important areas of functioning (1). The consequences of depressive disorders in terms of public health are vast. Global prevalence of depressive disorders is estimated in 322 million people, increasing 18.4% between 2005 and 2015. In addition, depression is ranked by WHO as the single largest contributor to global disability (7.5% of all years lived with disability in 2015); and a major contributor to suicide deaths, with an estimated close to 800 000 suicides per year (2,3).

Non-pharmacological therapies for depression include cognitive and behavioral therapy, electroconvulsive therapy (ECT), transcranial direct current stimulation (tDCS), transcranial magnetic stimulation (TMS), psychotherapy, physical activity and naturopathics, among others (6). In the last decade, innovative neuromodulation therapies have been developed. In this regard, Brain-Computer Interfaces based-neurofeedback helps to volitionally regulate brain activity through training the subject, thus providing self-administered therapy in neurorehabilitation (7,8). Up to date, non-invasive neurofeedback training have been built on different techniques, including electroencephalography (EEG), near-infrared spectroscopy (NIRS), magnetoencephalography (MEG) and functional magnetic resonance imaging (fMRI).

Functional MRI neurofeedback training can regulate brain activity and behavior; however, neurofeedback training is still on an early stage, and it remains unclear if those brain and behavioral changes can translate into clinical relevance (9–11). In addition, we aim to update neurofeedback knowledge, as the number of published studies is increasing every year and better quality of evidence is available. Finally, to our knowledge, this is the first systematic review assessing randomized control trials on fMRI neurofeedback training as a neurorehabilitation approach on depressive disorders. The main limitation of this systematic review might arise from the low number of extant RCTs.

## METHODS

### PROTOCOL AND REGISTRATION

This manuscript protocol complies with the ‘Preferred Reporting Items for Systematic reviews and Meta-Analyses’ (PRISMA) (12) guidelines for reporting systematic reviews and meta-analyses.

This systematic review will be submitted to registration on the International Prospective Register of Systematic Reviews (PROSPERO), https://www.crd.york.ac.uk/prospero/.

### ELIGIBILITY CRITERIA

#### Types of studies

We will only include randomized control trials.

#### Types of participants

We will include trials assessing participants clinically diagnosed with depressive disorder. Thus, we will include depressive disorders such as major depressive disorder, persistent depressive disorder (dysthymia), premenstrual dysphoric disorder, other specified depressive disorder, and unspecified depressive disorder, as defined by authors.

We will exclude clinical trials assessing substance/medication-induced depressive disorder, depressive disorder due to other medical conditions and disruptive mood dysregulation disorder, that can coexist with others depressive disorders, including major depressive disorder, attention-deficit/hyperactivity disorder, conduct disorder, and substance use disorders (1) in order to avoid confounding effects specially regarding overlaps in the region of interest when there are multiple disorders or different substance/medications involved.

We will exclude studies evaluating the effects on animal models.

#### Types of interventions

The intervention of interest is real-time fMRI neurofeedback training. We will not restrict our criteria to any number, frequency or duration of neurofeedback sessions. We will not restrict any feedback display (e.g. visual, auditory, tactile, proprioceptive) or feedback format (e.g. video clip, simple graphic, melody, tone). We will not restrict on how feedback was given (e.g. number of feedback sessions and runs, number of trainings, feedback strategy, feedback modality and content, amount of reward, strength of the magnetic field).

The comparison of interest is placebo (e.g. sham neurofeedback, neurofeedback from a largely unrelated brain signal, inversing the neurofeedback reward contingency), another active non-neurofeedback control (e.g. a similar type of computerized cognitive training, medication or standard therapy) or no treatment (neurofeedback plus standard treatment vs. standard treatment alone).

#### Types of outcomes

We will not consider outcomes as an inclusion criteria during the selection process. Any article meeting all the criteria except for the outcome criterion will be preliminarily included and evaluated in full text.

We will include the following outcomes according to recommendations by the ‘Consensus on the reporting and experimental design of clinical and cognitive-behavioural neurofeedback studies’ (CRED-nf) made for neurofeedback (13), and recommendations by the ‘Research Domain Criteria’ (RDoC) project proposed by the National Institute of mental Health (NIMH) made for neuroscience research (14,15).

- **Primary outcomes:**

1. Symptomatology/disease severity reduction.
- **Secondary outcomes:**

1. Quality of life.
2. Acceptability.
3. Adverse effects.
- **Other outcomes:**

1. Brain MRI metrics (e.g., BOLD signal percentage changes and individual differences on ‘upregulation’ and ‘downregulation’, DTI changes, cortical thickness changes).
2. Cognitive tasks (e.g., a working memory task).
3. Physiology measures defined as indices that do not necessarily tap circuits directly (e.g., heart rate, cortisol levels).

Primary and secondary outcomes will be presented at the ‘Summary of Findings table’ (SoFt).

### ELECTRONIC SEARCHES

We will conduct electronic searches for randomized controlled trials, without publication status or language restrictions in Pubmed, Embase and CENTRAL, Epistemonikos database, ClinicalTrials.gov and the International Clinical Trials Registry Platform (ICRTP).

The following search strategy was developed with the help of a librarian experienced in searching systematic reviews, using the combination of terms describing the intervention and the population of interest:

(((mood* OR affective*) AND (disorder* OR illness*)) OR dysthymi* OR depressi* OR unipolar* OR MDD OR ((premenstrual OR menstrual* OR “pre-menstrual") AND dysphori*) OR PMDD) AND ((“real-time functional MRI” OR “rt-fMRI-nf” OR “rtfMRI-nf” OR “functional-MRI” OR fMRI* OR rtfMRI* OR (functional* AND ("magnetic resonance” OR MRI*) AND (real-time OR “real time”)) OR ((BCI* OR BMI* OR “brain-computer” OR “brain-machine” OR “direct neural” OR (brain AND (computer* OR machine* OR interfac* OR regulat* OR “self-regulation”))))) AND (neurofeedback* OR “neuro feedback” OR “neuro-feedback” OR feedback))

We will adapt it to the syntax of each database.

### SEARCHING OTHER RESOURCES

We will review the reference lists of all included studies and other systematic reviews for additional potentially-eligible primary studies. We will also search in others sources as rtFIN database (http://www.rtfin.org/literature.html), review rtFIN conferences, Human Brain Mapping (OHBM) conferences, Society of Neuroscience (sfn) conferences from 2001 to date, and conduct a cross-citation search in Google Scholar using each included study as the index reference.

### SELECTION OF THE STUDIES

The results of the literature search will be incorporated into the screening software CollaboratronTM (16).

Two researchers will independently screen the titles and abstracts yielded by the search against the inclusion criteria. We will obtain the full reports for all titles that appear to meet the inclusion criteria or require further analysis to decide on their inclusion. Discrepancies between review authors will be resolved by discussion to reach consensus. If necessary, a third review author will be consulted to achieve a decision. We will record the reasons for excluding trials in any stage of the search and outline the study selection process in a PRISMA flow diagram adapted for the purpose of this project.

### EXTRACTION AND MANAGEMENT OF DATA

Two reviewers will independently conduct data extraction from each included study using the standardized methodology proposed by the Cochrane Collaboration Group (17). We will collect information regarding: the study design; the participant characteristics, including study eligibility criteria, diagnostic criteria, characteristics of participants at the beginning of the study (e.g. age, sex, comorbidity and disease severity); the intervention, including description of the intervention(s) and comparison intervention(s), ideally with sufficient detail for replication (e.g. data acquisition parameters, neurofeedback protocol, signal processing, and training characteristics); the outcomes (including measurement tool or instrument (e.g. characteristics of rating scales), specific metric used (e.g. percentage changes from baseline to a post-intervention time point, or post-intervention presence yes/no), method of aggregation (e.g. mean and standard deviation of scores in each group and timing of outcome measurements); the results; the source of funding; the conflicts of interest disclosed by the investigators and risk of bias assessment for each individual study. Discrepancies between review authors will be resolved by discussion to reach consensus. If necessary, a third review author will be consulted to achieve a decision.

### RISK OF BIAS ASSESSMENT IN INCLUDED STUDIES

The risk of bias for each randomized trial will be assessed using the first version of the Cochrane Collaboration tool for assessing risk of bias in randomized trials (18,19). Overall bias of each outcome studied will be determined according to the following six domains: selection bias, performance bias, detection bias, attrition bias, reporting bias, and other bias. The risk of bias will be evaluated as “low risk of bias”, “unclear risk of bias” or “high risk of bias” for each of these six domains. We will made a risk of bias table explaining the justifications for the allocation of the risk of bias. Discrepancies between review authors will be resolved by discussion to reach consensus. If necessary, a third review author will be consulted to achieve a decision.

### MEASUREMENTS OF TREATMENT EFFECT

For dichotomous outcomes, we will express the estimate of treatment effect of an intervention as risk ratios (RR) or odds ratios (OR) along with 95% confidence intervals (CI). For continuous outcomes, we will use mean difference and standard deviation (SD) to summarize the data, using a 95% CI. Whenever continuous outcomes are measured using different scales, the treatment effect will be expressed as a standardized mean difference (SMD) with 95% CI. When possible, we will multiply the SMD by a standard deviation that is representative from the pooled studies, for example, the SD from a well-known scale used by several of the studies included in the analysis on which the result is based. In cases where the minimally important difference (MID) is known, we will also present continuous outcomes as MID units or inform the results as the difference in the proportion of patients achieving a minimal important effect between intervention and control.

### DATA SYNTHESIS

If we include more than one trial we will conduct meta-analysis for studies clinically homogeneous using RevMan5 (20), using the inverse variance method with random effects model. For any outcomes where data was insufficient to calculate an effect estimate, a narrative synthesis will be presented.

### ASSESSMENT OF HETEROGENEITY

Initially this will be visually evaluated by means of forest plot. Since statistical heterogeneity is inevitable, inconsistency between the studies will be quantified using the I2 statistic. The latter describes the percentage of variability in the effect estimates that is due to heterogeneity rather than sampling error (chance). The interpretation of the I2 statistic will be carried out as follows: 0% to 40%: it may not be important; 30% to 60%: it may represent moderate heterogeneity; 50% to 90%: it may represent significant heterogeneity; 75% to 100%: considerable heterogeneity (21). If the calculation of the I2 statistic indicates heterogeneity between the results of the studies, the possible causes will be analyzed and a meta-analysis will be discussed or not.

### SUBGROUP AND SENSITIVITY ANALYSIS

We will conduct our main analyses combining all interventions and populations, but when possible we will assessed specific subgroup analyses to explore possible sources of heterogeneity:

- Age (e.g. <18 years old, ≥ 18 years old)
- Type of depressive disorder
- Severity of the depressive disorder at baseline
- Number of feedback sessions and runs
- Number of trainings
- Feedback strategy
- Feedback modality and content
- Amount of reward
- Strength of the magnetic field

We will perform sensitivity analysis excluding high risk of bias studies. In cases where the primary analysis effect estimates and the sensitivity analysis effect estimates significantly differ we will either present the low risk of bias (adjusted sensitivity analysis estimates) or present the primary analysis estimates but downgrading the certainty of the evidence because of risk of bias.

We will also consider for sensitivity analysis different diagnostic criteria for depression disorders (DSM-5 versus other criteria used by authors).

### ASSESSMENT OF CERTAINTY OF EVIDENCE

The certainty of the evidence for primary and secondary outcomes will be judged using the Grading of Recommendations Assessment, Development and Evaluation working group methodology (GRADE Working Group). For the main comparisons and primary and secondary outcomes, we will prepare a Summary of Findings (SoF) table. Other outcomes will be presented as an appendix.

### ETHICS AND DISSEMINATION

This protocol was presented for dispensation to the scientific ethics committee of ‘Pontificia Universidad Católica de Chile’. This review will only include published data that already received ethical approval before publication, and researchers will not access information that could lead to the identification of an individual participant. The results of these articles will be widely disseminated via peer-reviewed publications, social networks, and will be sent to relevant organizations discussing this topic.

### FUNDING

This review was not commissioned by any organization and did not receive external funding.

## Data Availability

does not apply

